# Left frontal white matter links to rhythmic processing relevant to speech production

**DOI:** 10.1101/19013458

**Authors:** Rose Bruffaerts, Jolien Schaeverbeke, Manon Grube, Silvy Gabel, An-Sofie De Weer, Eva Dries, Karen Van Bouwel, Timothy D Griffiths, Stefan Sunaert, Rik Vandenberghe

## Abstract

Recent evidence suggests that a shared neurocomputational mechanism tracks the temporal structure of auditory input and speech output. We investigate the link between speech production and rhythmic abilities in 24 healthy controls and 23 patients with primary progressive aphasia, including 12 non-fluent variant patients (NFV) with apraxia of speech. Seventy-five percent of NFV cases displayed poorer rhythmic abilities. Rhythmic abilities and speech rhythm were significantly correlated in both controls and patients. Deformation-based morphometry and diffusion tensor imaging were used to identify white matter atrophy related to impaired rhythmic abilities in NFV. Left frontal white matter volume adjacent to the supplementary motor area (SMA) correlated with rhythmic abilities. Metrics of the left Aslant tract, which is typically damaged in NFV cases, also correlated with rhythmic abilities. Our results support the existence of a common temporal scaffolding mechanism structuring perceptual input and speech output. This mechanism partly explains abnormal speech in NFV.

## Introduction

Recent mechanistic models argue for a key role of rhythm processing in both speech production and speech perception (Poeppel and Assaneo, 2020). Here, we examine this link in individuals with the non-fluent variant (NFV) of primary progressive aphasia (PPA) who suffer from apraxia of speech (AOS), hampering speech production. The rhythmicity of speech originates at the suprasegmental level, i.e. the fairly regular production of syllables. Across natural languages and speakers, the suprasegmental temporal structure of speech varies within similar frequency bands (2-10 Hz) (Ding et al., 2017). Speech rhythm also enhances perception: speech perception is optimal between 2 and 8 Hz (Ghitza and Greenberg, 2009) and the auditory cortex is tuned to these frequencies (Boemio et al., 2005). The relationship between the neural processes supporting speech rhythm and speech perception is under debate. Grube et al. (2012) have proposed a “temporal scaffolding mechanism”, a common neurocomputational mechanism which structures both perceptual input and perceptual output, i.e. speech, in time. For instance, they observed that the ability to discriminate rhythm correlated with phonological abilities (reading, repetition) in adolescents. Patients with NFV represent a unique opportunity to test this temporal scaffolding hypothesis. Clinically, NFV patients do not have consistent problems producing individual phonemes, but have difficulties with the suprasegmental timing of their speech resulting in an abnormal speech rhythm (Duffy et al., 2017). At the auditory perceptual level, abnormalities have been documented in NFV (Goll et al., 2010; Warren et al., 2005). Our prior work demonstrated that NFV patients’ rhythmic abilities were poorer compared to controls and patients with the semantic variant (SV) of PPA (Grube et al., 2016).

By determining rhythmic abilities in an extended PPA cohort including 12 patients with NFV (Table 1) and 11 patients with SV, we here investigate two predictions stemming from the temporal scaffolding hypothesis. First, our prior work focused on the impairment of rhythmic abilities in NFV at the group level. Here, we hypothesize that speech rhythm will correlate to rhythmic abilities at the individual level and we test this on connected speech samples of both controls and patients. Second, we hypothesize that neuroimaging of the NFV group may enable us to identify the neuroanatomical correlate of the temporal scaffolding mechanism. Grey matter atrophy is consistently found in NFV in the left opercular part (BA44) of the inferior frontal gyrus (IFG), insula, premotor and the supplementary motor areas (SMA) (Gorno-Tempini et al., 2011; Rogalski et al., 2011). The degree of atrophy in these regions correlates with the severity of speech rhythm abnormalities in NFV (Ballard et al., 2014). SMA has been identified as a gray matter correlate of auditory temporal regularity detection in healthy volunteers (Grahn and Brett, 2007; Grahn and Schuit, 2012) and NFV (Hardy et al., 2017b, 2017a). The correlational neuroimaging findings in NFV align perfectly with recent neurophysiological evidence. For instance, oscillatory activity in IFG and motor regions has been linked to speech perception (Assaneo and Poeppel, 2018; Keitel et al., 2018; Magrassi et al., 2015). Cooling of these regions leads to disruption of speech rhythm and dysarthria, respectively (Long et al., 2016).

**Table 1.**
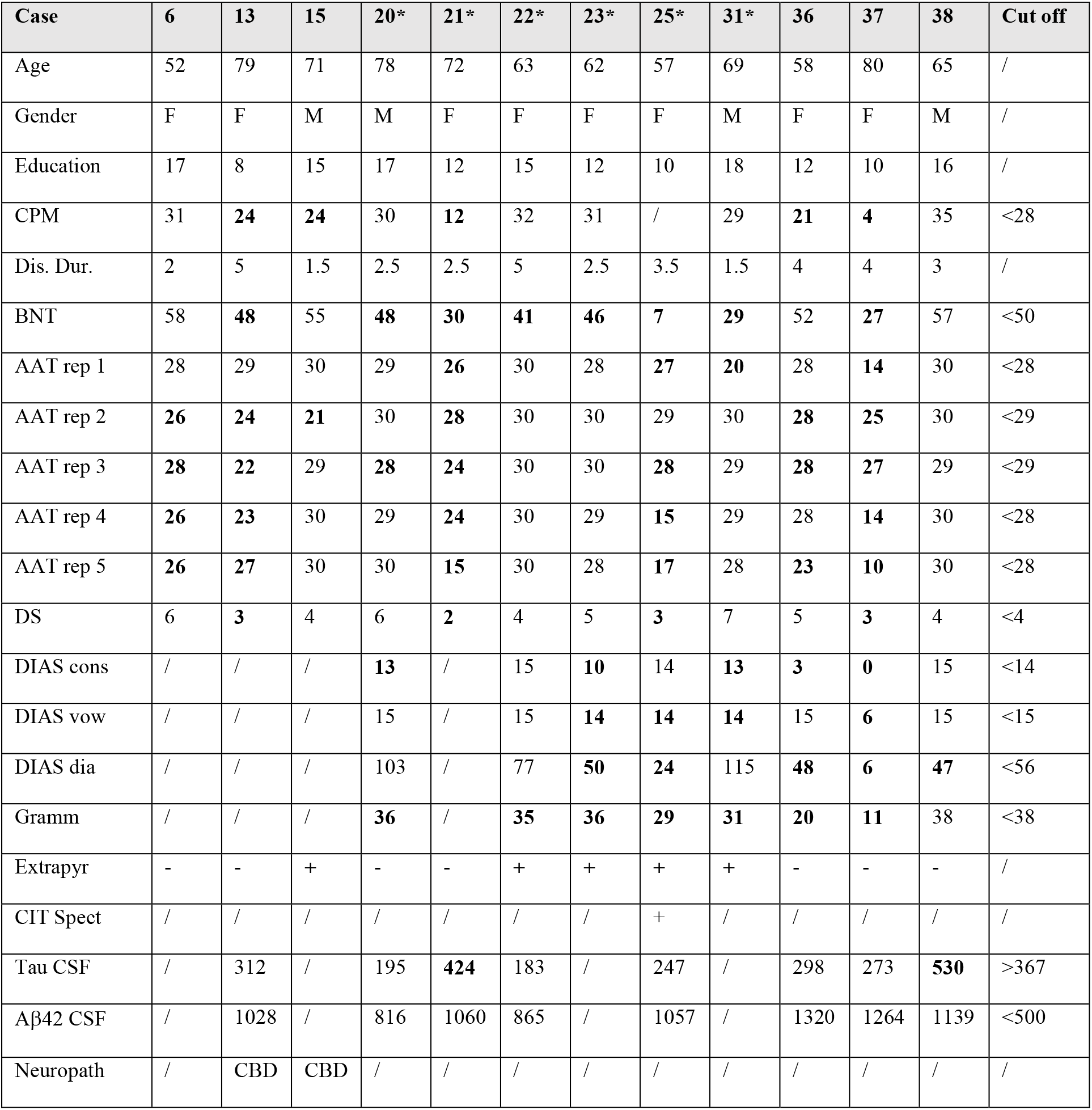
Characteristics of NFV patients. Norms were calculated as 2 standard deviations below the mean in the age- and education-matched group of healthy controls (n=29). CPM: Raven’s progressive matrices, Dis. Dur.: disease duration (years), BNT: Boston Naming Test, AAT: Akense Afasie Test repetition scores part 1 to 5, DS: digit span forward, DIAS cons: DIAS repetition of consonants, DIAS vow: DIAS repetition of vowels, DIAS dia: DIAS diadochokinesis score, Gramm: grammaticality score WEZT, Extrapyr: extrapyramidal signs upon examination, Neuropath: anatomopathological findings, * Mixed variant of PPA.

| Case | 6 | 13 | 15 | 20* | 21* | 22* | 23* | 25* | 31* | 36 | 37 | 38 | Cut off |
| --- | --- | --- | --- | --- | --- | --- | --- | --- | --- | --- | --- | --- | --- |
| Age | 52 | 79 | 71 | 78 | 72 | 63 | 62 | 57 | 69 | 58 | 80 | 65 | / |
| Gender | F | F | M | M | F | F | F | F | M | F | F | M | / |
| Education | 17 | 8 | 15 | 17 | 12 | 15 | 12 | 10 | 18 | 12 | 10 | 16 | / |
| CPM | 31 | **24** | **24** | 30 | **12** | 32 | 31 | / | 29 | **21** | **4** | 35 | <28 |
| Dis. Dur. | 2 | 5 | 1.5 | 2.5 | 2.5 | 5 | 2.5 | 3.5 | 1.5 | 4 | 4 | 3 | / |
| BNT | 58 | **48** | 55 | **48** | **30** | **41** | **46** | **7** | **29** | 52 | **27** | 57 | <50 |
| AAT rep 1 | 28 | 29 | 30 | 29 | **26** | 30 | 28 | **27** | **20** | 28 | **14** | 30 | <28 |
| AAT rep 2 | **26** | **24** | **21** | 30 | **28** | 30 | 30 | 29 | 30 | **28** | **25** | 30 | <29 |
| AAT rep 3 | **28** | **22** | 29 | **28** | **24** | 30 | 30 | **28** | 29 | **28** | **27** | 29 | <29 |
| AAT rep 4 | **26** | **23** | 30 | 29 | **24** | 30 | 29 | **15** | 29 | 28 | **14** | 30 | <28 |
| AAT rep 5 | **26** | **27** | 30 | 30 | **15** | 30 | 28 | **17** | 28 | **23** | **10** | 30 | <28 |
| DS | 6 | **3** | 4 | 6 | **2** | 4 | 5 | **3** | 7 | 5 | **3** | 4 | <4 |
| DIAS cons | / | / | / | **13** | / | 15 | **10** | 14 | **13** | **3** | **0** | 15 | <14 |
| DIAS vow | / | / | / | 15 | / | 15 | **14** | **14** | **14** | 15 | **6** | 15 | <15 |
| DIAS dia | / | / | / | 103 | / | 77 | **50** | **24** | 115 | **48** | **6** | **47** | <56 |
| Gramm | / | / | / | **36** | / | **35** | **36** | **29** | **31** | **20** | **11** | 38 | <38 |
| Extrapyr | - | - | + | - | - | + | + | + | + | - | - | - | / |
| CIT Spect | / | / | / | / | / | / | / | + | / | / | / | / | / |
| Tau CSF | / | 312 | / | 195 | **424** | 183 | / | 247 | / | 298 | 273 | **530** | >367 |
| Aβ42 CSF | / | 1028 | / | 816 | 1060 | 865 | / | 1057 | / | 1320 | 1264 | 1139 | <500 |
| Neuropath | / | CBD | CBD | / | / | / | / | / | / | / | / | / | / |

White matter changes also contribute to impaired speech production in NFV (Canu et al., 2019; Galantucci et al., 2011; Mandelli et al., 2014), which motivated our choice to investigate whether white matter changes correlate to rhythmic abilities in NFV. We used two independent modalities: tensor-based deformations of the brain (deformation-based morphometry, DBM) and diffusion tensor imaging (DTI). We opted for DBM rather than voxel-based morphometry (VBM) because automated segmentation in regions of abnormal grey and white matter might be unreliable and because DBM allows visualization of changes in subcortical structures containing grey and white matter (Cardenas et al., 2007). DBM is also easier to interpret than VBM since it reflects atrophy without inference from other pathological white matter changes. Furthermore, we complement DBM with DTI. DTI is sensitive to white matter damage even at the individual level (Sajjadi et al., 2013), which is beneficial given the relatively rarity of NFV. Damage of the left Aslant tract, which connects BA44 to medial frontal areas including the SMA (Catani et al., 2013), is considered specific for the NFV phenotype (Canu et al., 2019; Mandelli et al., 2014). We thus focused our DTI analysis on the left frontal Aslant tract.

In summary, we here examine the link between speech production and rhythmic abilities to strengthen the evidence for a neurocomputational temporal scaffolding mechanism and to understand the neurobiological mechanism underpinning impaired speech production in NFV.

## Results

### Psychoacoustic tasks

A total of 47 participants performed a psychoacoustic test battery: 12 NFV (Table 1), 11 SV and 24 healthy mature controls. The test battery consisted of four tasks determining each participant’s ability to perceive the timing of meaningless auditory stimuli. Two tasks (single time-interval duration discrimination (r1), isochrony deviation detection (r2), Fig 1A, supplementary audio samples) probed the detection of local deviations of temporal structure. Two tasks (metrical pattern discrimination tasks using a strongly (r3) or weakly (r4) metrical reference, Fig 1A, supplementary audio samples) probed the detection of higher-order/suprasegmental deviations of temporal structure. The latter tasks (r3,r4) measure the “rhythmic abilities” of each participant. For each task and each participant, a threshold of discrimination is obtained from an adaptive staircase procedure, with higher thresholds equaling poorer abilities to detect deviations. Based on the suprasegmental abnormalities found in the patients’ speech, we postulate that the tasks indexing suprasegmental timing (r3,r4) will be most impaired in NFV.

**Figure 1:**
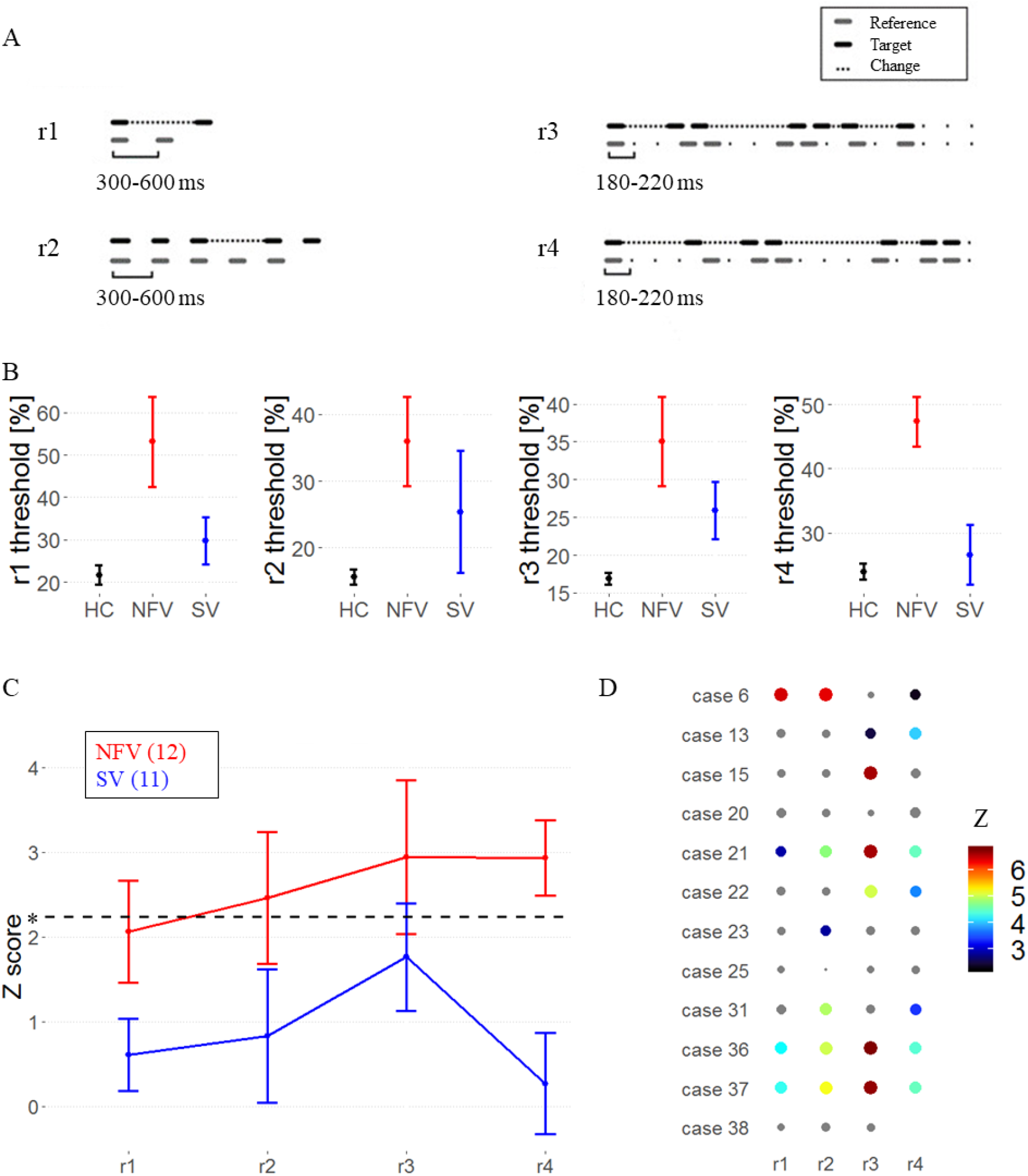
Psychoacoustic tasks A) Experimental design, audio samples in supplemental. B) Mean raw thresholds (and standard error of the mean) across controls, NFV and SV patients. C) Mean Z scores (and standard error of the mean) across PPA subtypes. Dotted line represents the Z cut-off for Bonferroni-corrected P<0.05. D) Z-scores of rhythm tests in NFV (case numbers refer to table 1). The size and color of dots reflect z-score value, non-significant values are gray. Missing data indicates that the patient was unable to perform the task.

In a group-level analysis, performance on the psychoacoustic tasks was poorer in NFV compared to controls (Fig 1AB): mean Z scores were above the threshold (P<0.05 Bonferroni-corrected) in NFV for discrimination of strongly metrical sequences (r3, mean Z: 2.94), discrimination of weakly metrical sequences (r4, mean: 2.93) and isochrony deviation detection (r2, mean: 2.46) (Fig 1B). We compared the test scores between the NFV and SV. This resulted in significantly poorer scores in NFV for the discrimination of weakly metrical sequences (r4, P = 0.001, Hedges’ g: 1.48) (Fig 1BC). At the individual level, deficits were observed mainly in NFV patients (Z > 2.24) (Fig 1D). The weakly metrical pattern discrimination task (r4) demonstrated a significant impairment in 7 NFV (Fig 1D) and 2 SV patients. Similarly, strongly metrical pattern discrimination (r3) was impaired in 6 NFV (Fig 1D) and 4 SV, as well as isochrony deviation detection (r2) in 6 NFV and 2 SV patients. Single time-interval discrimination (r1) was impaired in just 4 NFV and 1 SV patients. In summary, 75% of NFV cases were impaired in one or more of the tasks and 36.4% of SV cases.

### Correlation of psychoacoustic tasks with speech rhythm & rate

The pairwise variability index (PVI) was calculated on a 2-minute audio sample (“Cookie Theft Scene” description) as a measure of speech rhythm (Ballard et al., 2014; Duffy et al., 2017). PVI reflects the relative duration of the stressed versus unstressed syllable and was calculated for polysyllabic words with strong-weak pattern (stress on the first syllable) and a weak-strong pattern (stress on the second syllable). PVI values were closer to zero (relatively equal stress) for words with a weak-strong stress pattern in NFV compared to controls and SV (one-way ANOVA F(2,35)=5.37, P = 0.009, Fig 2A). No significant between-group differences were found for strong-weak words. The speech rate (defined as utterances per minute) in NFV was significantly slower (one-way ANOVA F(2,35)=20.39, P < 0.001, Fig 2B). At the individual level, 40% of NFV and 10% of SV displayed changes in the speech rhythm for weak-strong words and 30% of NFV patients for strong-weak words. In the NFV group, 70% of individuals had a significantly slowed speech rate compared to controls.

**Figure 2:**
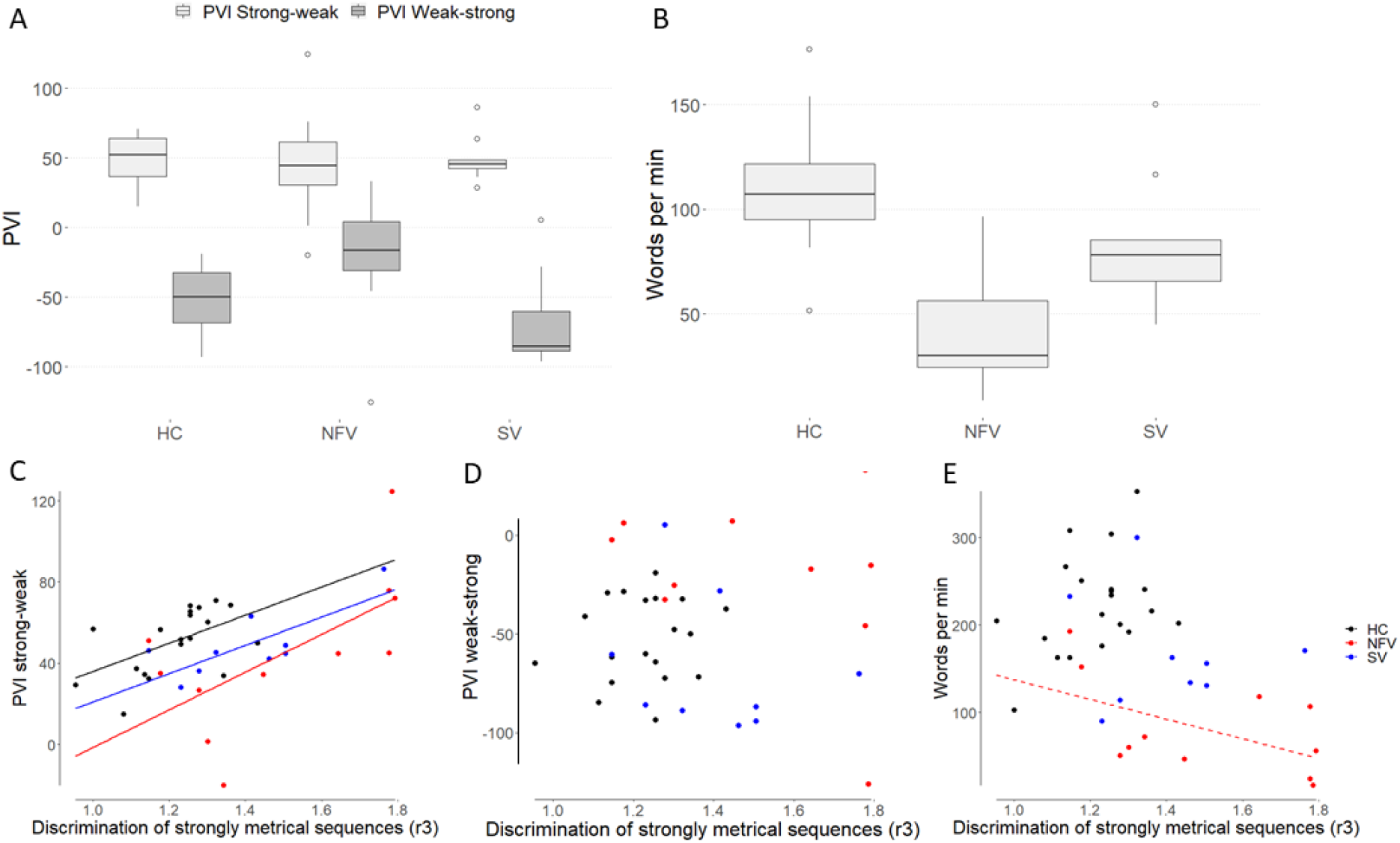
Speech rhythm & rate. A) Median PVI values for strong-weak and weak-strong words in the 3 participant groups. B) Median speech rate. C) Correlation of PVI for strong-weak words and strong metric sequences threshold (r3, log-transformed), regression line indicates a significant correlation at the subgroup level, D) Correlation of PVI for weak-strong words and strong metric sequences threshold (r3, log-transformed) E) Correlation of PVI for strong-weak words and strong metric sequences threshold (r3, log-transformed), dashed regression line indicates a trend at the subgroup level for NFV.

Across all participants, PVI for strong-weak words correlated with strongly metrical pattern discrimination (r3) (R = 0.444, P = 0.004, Fig 2C). In an analysis per subgroup, this correlation was found for NFV (R = 0.634, P = 0.036), SV (R = 0.761, P = 0.017) and controls (R = 0.531, P = 0.016). This means that participants with poorer rhythmic abilities, displayed greater duration differences between the first and second vowels of words with a strong-weak stress pattern. No correlation was found with any of the other psychoacoustic tasks (r1,r2,r4) and no correlation was found with the PVI for weak-strong words (all P>0.1). Strongly metrical pattern discrimination (r3) also correlated with speech rate at across the whole sample (R = -0.472, P = 0.002, Fig 2E): rhythmic abilities were poorer when speech was slower. A trend for significance was observed in the NFV subgroup (R = -0.538, P = 0.088), but this correlation was absent in the other subgroups (SV: R = -0.139, P = 0.722; controls: R = 0.354, P = 0.126).

### White matter changes: deformation-based morphometry

The expected atrophy patterns per PPA subgroup were observed using deformation-based morphometry (Fig 3). In the group consisting of 12 NFV cases, atrophy was observed mainly in the frontal lobes compared to controls, with a left-sided predominance (Fig 3AB). In the SV group, atrophy was localized to the anterior temporal lobes (Fig 3A). In the NFV group, voxel-wise multiple linear regression showed that the strongly metrical rhythm discrimination task (r3) negatively correlated with changes in the deformation field in the left frontal white matter (MNI = -20,20,-36; -17,8,48; -9,39,50; kE 2426 voxels, Z score: 4.92) (Fig 4AB). This negative correlation indicates that poorer rhythmic abilities (i.e. larger thresholds) were linked to more atrophy. For illustrative purposes, we plotted the individual NFV thresholds for the strongly metrical discrimination task (r3) versus volume loss in this region (R = -0.316, R^2^ = 0.100, P < 0.001) (Fig 4C). DBM analysis yielded no other significant correlations with the psychoacoustic tasks in the NFV or SV subgroups.

**Figure 3:**
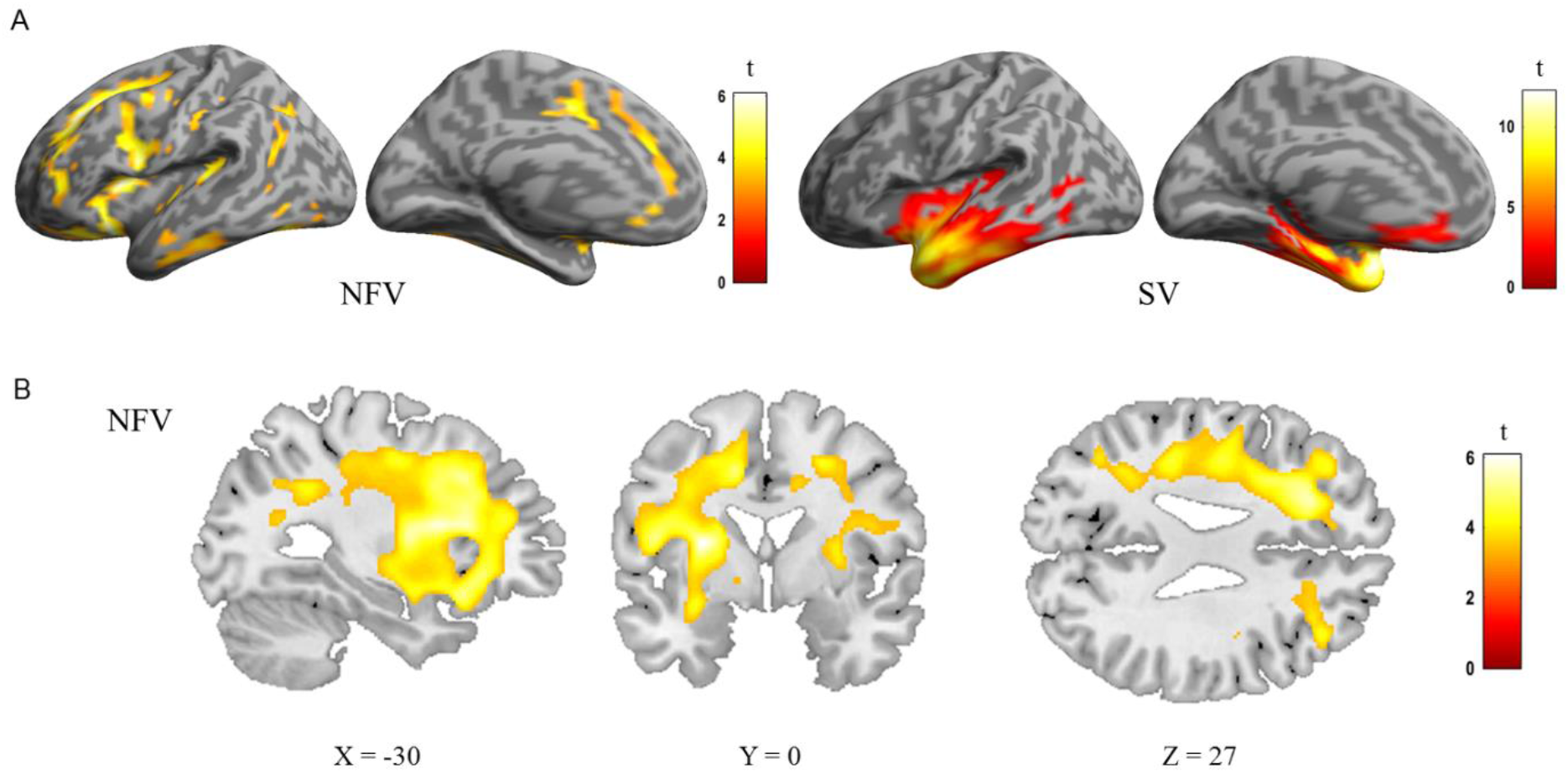
DBM analysis: comparison of controls, the NFV and SV patients. A) Renderings shows deformation of 12 NFV and 11 SV compared to 24 controls (cluster-level FWE-corrected P<0.05). B) Slices in NFV

**Figure 4:**
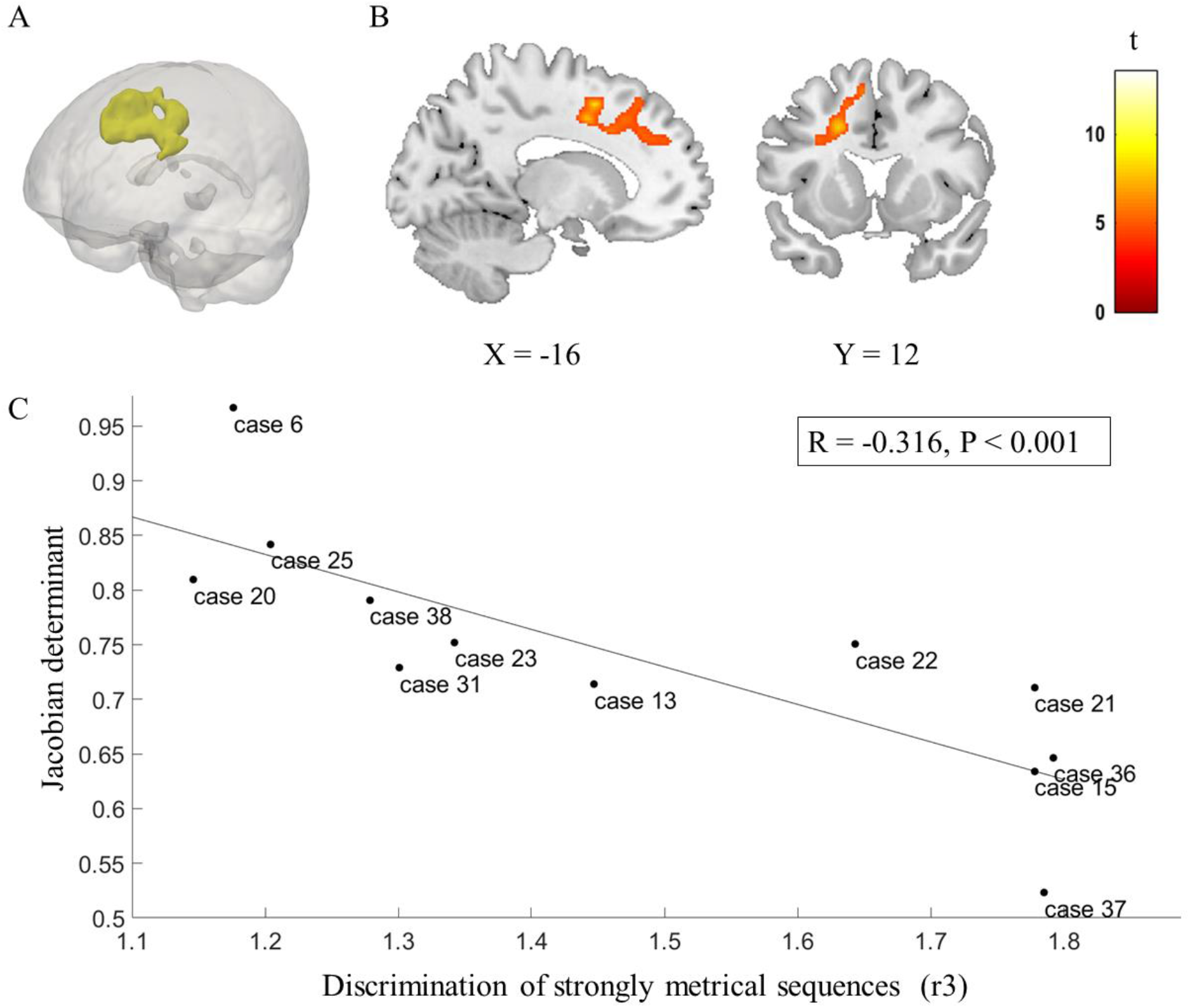
DBM of strongly metrical sequence in NFV patients. A) Group rendering B) Slices C) Correlation between volume loss and strongly metrical sequence thresholds (r3, log-transformed) in NFV patients in the region of interest (AB), exploratory plot for illustrative purposes (case numbers refer to table 1)

### White matter changes: Diffusion Tensor Imaging

We performed DTI in a subset of 7 NFV, 7 SV and 20 controls to obtain an independent white matter measure. A comparison between NFV, SV and controls showed reduced FA in NFV in the left inferior frontal region, the corpus callosum and the anterior cingulate (Fig 5A). MD was widely increased in NFV, with a predominance in both frontal lobes (Fig 5B). In SV, FA was reduced and MD was increased in both anterior temporal lobes (not shown). We then compared FA and MD between NFV and SV specifically within the template of the left Aslant tract derived from the controls. Although FA was similar between NFV and SV (P=0.175, Hedges’ g: -0.72, Fig 5C), MD was higher in NFV compared to SV (P = 0.038, Hedges’ g: 1.17, Fig 5D). In NFV, MD in the left Aslant tract was increased when the performance on the strongly metrical rhythm discrimination task was weaker (r3) (R = 0.815, R^2^ = 0.664, P = 0.026). Although this was not significant, a trend was observed with FA (R = - 0.708, R^2^ = 0.501, P = 0.075). Neither FA nor MD in the left Aslant tract correlated with performance on any other task in NFV (r1,r2,r4, all P>0.231). Visual inspection of the left Aslant tract in NFV showed that this tract overlapped with the region where there were white matter volume changes identified by DBM (Fig 5E).

**Figure 5:**
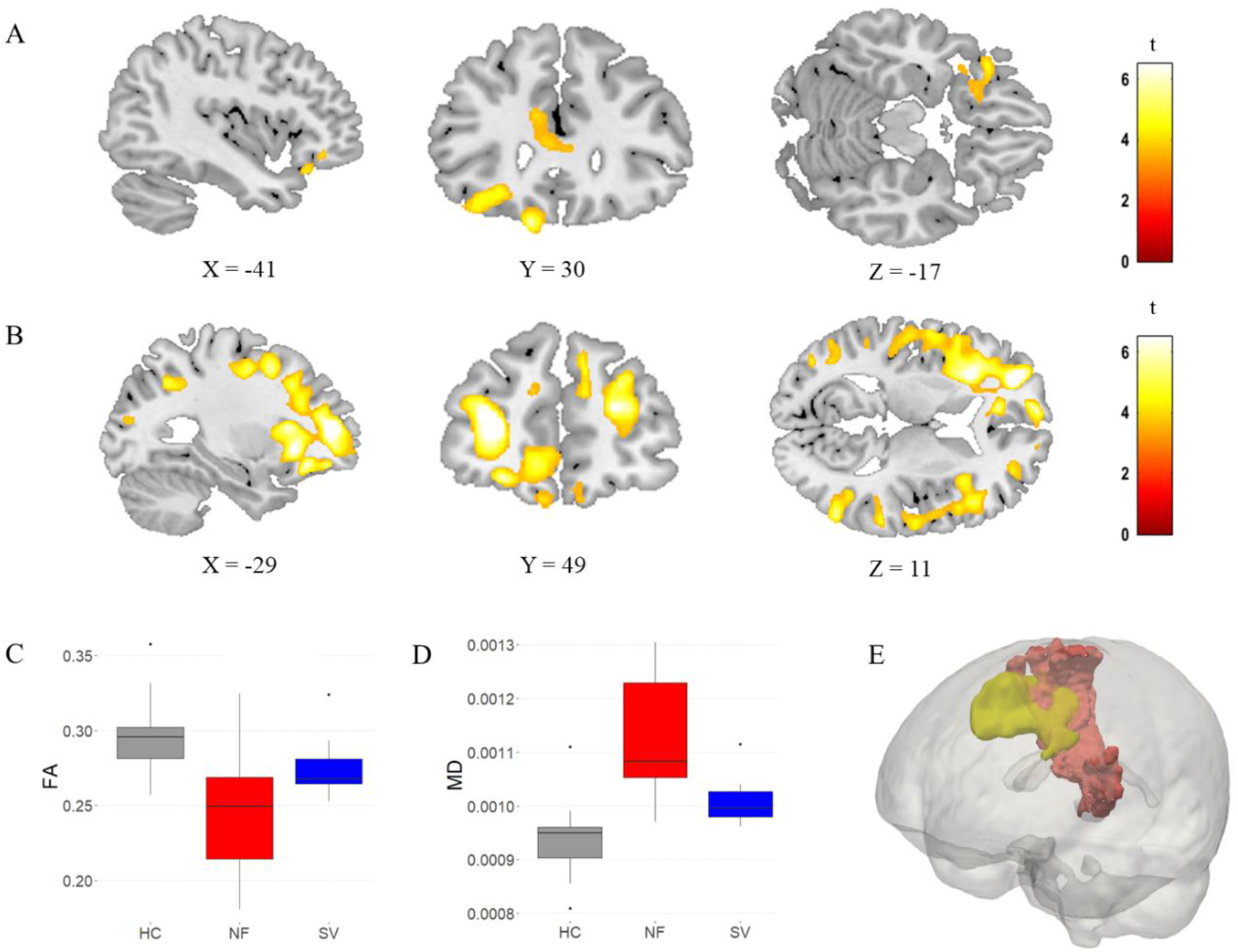
DTI Metrics. A) FA in NFV patients versus controls (cluster-level FWE-corrected P<0.05). B) MD in NFV cases versus controls (cluster-level FWE-corrected P<0.05). Comparison of C) FA and D) MD in the left frontal Aslant tract between the NFV and SV subtypes and healthy controls (HC). E) Visualization of the Aslant tract in NFV, based on the 75% overlap of individual tracts in NFV (red) as well as the region of interest derived from DBM (yellow)

## Discussion

In patients with impaired speech production due to NFV, we investigated whether speech rhythm and rhythmic abilities are linked. Such a link, indicative of a common neurocomputational temporal structuring mechanism, was predicted by the temporal scaffolding hypothesis (Grube et al., 2016, 2012). In an extended cohort of 12 NFV, we confirmed that rhythmic abilities are overall poorer compared to controls. Behaviorally, we observed a correlation between rhythmic abilities and speech rhythm for NFV as well as for SV and healthy mature individuals, in agreement with a coupling between auditory perception and speech production. Based on our experiments which make use of meaningless acoustic stimuli, we argue that the role of temporal scaffolding might well extend beyond the linguistic domain. In the NFV group, DBM demonstrated that atrophy in the left frontal lobe correlated with the patients’ rhythmic abilities. We complemented DBM with DTI to provide an independent measure of white matter changes. DTI confirmed a correlation between damage to the left Aslant tract and rhythmic abilities in NFV. Given the prior work implicating the left Aslant tract to motor speech production deficits in NFV (Canu et al., 2019; Catani et al., 2013; Mandelli et al., 2014), our results suggest it is a relevant (part of a) common anatomical substrate for impaired rhythmic abilities and speech production. Whilst our findings are correlational, the results for speech rhythm complement the two independent white matter metrics. This strengthens the evidence base for a well-defined neurocomputational mechanism of temporal scaffolding linking perception and speech production. In NFV, the concept of impaired temporal scaffolding provides insight into temporal irregularities in spontaneous speech, as well as the role of left frontal regions in this process. Moreover, the link between speech rhythm and rhythmic abilities in all subgroups suggests that left frontal white matter may also play a role in structuring perceptual input and speech output in time in the healthy brain.

Although four psychoacoustic tasks were performed, both speech rhythm and left frontal lobe atrophy in NFV were linked specifically to impaired performance on the strongly metrical rhythm discrimination task (r3). This task is conceptually different from the single time-interval duration discrimination task (r1) and the isochrony deviation detection task (r2): determining the metricality of a tone sequence (r3) requires processing of the higher-order temporal structure determined by the grouping of salvos of notes that induce the sense of a regularly occurring metrical ‘beat’ (Grube and Griffiths, 2009). Metricality-based rhythm discrimination (r3) necessitates detecting global deviations distributed across the entire sequence. The correlation between rhythmic abilities and speech rhythm may stem from the common processes required to integrate the higher-order/suprasegmental temporal structure. Our results are in agreement with prior work in PPA that demonstrates the detection of temporal changes between syllables was more impaired compared to controls when stimuli contained a higher number of syllables (Rohrer et al., 2012). In contrast, the single time-interval duration discrimination task (r1) and the isochrony deviation detection task (r2) test lower-order differences in timing in a simple isochronous sequence based on a local deviation. The weakly metrical rhythm discrimination task (r4) is more challenging as it does not rely on a clear metrical beat (Grube and Griffiths, 2009) (higher thresholds for r4 versus r3 in controls, P<0.001). Perhaps more domain-general processes play a role in this task, but additional manipulations are required to confirm this hypothesis. Keeping in mind the labor-intensive administration of our tasks, we reached a considerable albeit modest sample size. Further validation requires a larger multicentric sample given the relative rarity of PPA.

Interestingly, we observed a coupling between speech rhythm and rhythmic abilities in all participants groups. Such a coupling aligns with the temporal scaffolding hypothesis because it supports the idea that both speech production and perception tap into a common neural mechanism sustaining rhythmic processing. The fact that the coupling even holds in patients groups such as NFV with AOS further evidences this claim. Our behavioral results complement the existing literature on speech rhythmicity. First, speech rhythm, measuring the relative lengthening of a stressed versus unstressed vowel, can be viewed a synchronicity measure. Interindividual differences in the tendency to synchronize spoken syllables to auditory input, have been ascribed to a similar coupling mechanism between perception and production localized to the left IFG and white matter neighboring the auditory cortex (Assaneo et al., 2019). Second, another proposed mechanism for the coupling between speech perception and production is that the motor cortex contains predictions about the speaker’s vocal output which are updated (Liebenthal and Möttönen, 2018). This loop can be manipulated using “delayed auditory feedback”, an experimental procedure in which the auditory feedback of the speaker’s voice is delayed. This manipulation results in an individual-specific degree of speech perturbation in healthy speakers (Chon et al., 2013). Surprisingly, this manipulation improves speech production in some NFV patients (Hardy et al., 2018), indicative of changes in the neural substrate involved in temporally structuring input and output. However, our behavioral results cannot explain the complete extent of speech rhythm abnormalities in NFV with AOS. In accordance with prior work (Ballard et al., 2014; Duffy et al., 2017), we observed that speech rhythm of weak-strong words is most frequently impacted in NFV. Because coupling of rhythmic abilities and speech rhythm for weak-strong words was absent in all participants group, we currently cannot provide a mechanistic account for these speech rhythm abnormalities in NFV with AOS.

Using two independent white matter metrics, we identified an overlapping white matter substrate in the left frontal lobe which may play a role in both rhythmic abilities and speech production and is thus a candidate component of the temporal scaffolding mechanism. We observed white matter degeneration in the left frontal Aslant tract close to the SMA. The Aslant tract connects the superior frontal gyrus/SMA to the IFG, cortical regions that have previously been implicated in rhythmic processing (Poeppel and Assaneo, 2020). SMA was linked to temporal regularity discrimination (Grahn and Schuit, 2012; Hardy et al., 2017a) and apraxia of speech (Tetzloff et al., 2018; Whitwell et al., 2013) in NFV. IFG plays a role in speech rhythm (Long et al., 2016) and synchronizing speech to external auditory stimuli (Assaneo et al., 2019). The correlation between rhythmic abilities and left Aslant tract metrics align with the contemporary view that speech rhythm production and perception are sustained by a left hemispheric network rather than a single cortical region (Mandelli et al., 2016; Poeppel and Assaneo, 2020). Our focus on the left Aslant tract, motivated by our choice of study population, does not preclude that other white matter tracts e.g. tracts connecting to the auditory cortices (Assaneo et al., 2019), may also play an important role in this network. Accordingly, the DBM analysis also demonstrated that the region in which atrophy correlated to rhythmic abilities extends beyond the boundaries of the left Aslant tract.

One consideration is whether the white matter changes reflect tau pathology, which is found in up to 88% of NFV patients (Spinelli et al., 2017). In particular, DTI metrics have been put forward as a marker of tauopathy and other proteinopathies (Downey et al., 2015; Mahoney et al., 2013). DTI imaging is sensitive to changes caused by tau pathology at the single-subject level (Sajjadi et al., 2013), presumably because of underlying glial pathology (Forman et al., 2002), e.g. by myelin injury or changes in other structures that affect water diffusion (Galantucci et al., 2011). Here, we performed DTI imaging in a small subset of the study population (7 NFV, 7 SV and 20 controls) and because of the small sample size, we interpret these findings with appropriate caution. Our results are in alignment with prior work in PPA (Powers et al., 2013): MD changes were more pronounced than FA changes in NFV patients (Whitwell et al., 2010). In our study, neuropathological data is lacking for most patients, thus prohibiting us from making strong claims in relation to pathology. We would not advocate linking tauopathy to a simple DTI parameter. Rather, our findings advance the broader characterization of the possible disease-specific involvement of white matter tracts. Our results align with the “molecular nexopathy” paradigm (Warren et al., 2013): the left frontal network containing IFG and SMA as nodes demonstrate a selective vulnerability to tau protein, which could spread locally through the left Aslant tract in a prionlike fashion. Even if certain proteinopathies are strongly linked to predictable phenotypes of network disruption, the molecular nexopathy paradigm does not propose complete specificity. Accordingly, we also observed impaired rhythmic abilities in some SV patients. As the disease progresses, PPA subtypes exhibit convergence of their atrophy patterns (Bruffaerts et al., 2020; Leyton et al., 2019), even though the underlying neuropathology is different. This convergence of atrophy patterns may explain that some SV demonstrated impaired rhythmic abilities. Finally, we acknowledge that even though all our NFV cases exhibited clear AOS, the clinical picture often included other symptoms typically observed in NFV such as agrammatism or extrapyramidal signs and we cannot quantify how these findings relate to the observed white matter changes. Studying a related phenotype with isolated AOS, primary progressive AOS (ppAOS) (Josephs et al., 2012), would aid in this discrimination, but this phenotype is even more rare than NFV (Duffy et al., 2020). Neuroimaging analyses of patients with ppAOS are in agreement with our results: reduced FA was observed in the left SMA in ppAOS (Utianski et al., 2018) and functional connectivity analysis demonstrated that SMA is disconnected from the speech and language network in ppAOS (Botha et al., 2018).

In summary, we observed a coupling between speech rhythm and rhythm perception in controls and patients with impaired speech production, further supporting the notion of a common temporal scaffolding mechanism structuring input and output. Temporal scaffolding abnormalities explain part of the speech rhythm abnormalities in NFV. Our DBM and DTI findings in NFV showed concordant evidence that rhythmic abilities correlate with left frontal white matter atrophy, overlapping with the substrate for impaired speech production. Overall, we provided novel evidence for a common neurocomputational mechanism for speech rhythm and rhythm perception which elucidates speech abnormalities and which may contribute to the development of tailored rehabilitation strategies.

## Methods

### Participants

The study was approved by the Ethics Committee, University Hospitals Leuven. All participants provided written informed consent in accordance with the Declaration of Helsinki. PPA patients were recruited via the memory clinic University Hospitals Leuven. A consecutive series of 37 patients who fulfilled the international consensus criteria for PPA (Gorno-Tempini et al., 2011) enrolled for the experiment (2011-2019). The first 23 patients were described in (Grube et al., 2016), and the same case numbers are used. Six patients were excluded due to: hearing loss (n = 3); lack of ability to perform the experimental tasks due to disease severity (n = 2); lack of cooperation (n = 1). The remaining 31 patients were able to undergo the extensive testing and produce reliable data. Before enrollment, each patient was classified according to the 2011 recommendations (Gorno-Tempini et al., 2011). The classification relied on the clinical evaluation by an experienced neurologist (R.V.). Twelve cases were classified as NFV (Table 1), 11 as the semantic variant (SV), and 8 as the logopenic variant (LV). The LV group will not be discussed because of the smaller sample size compared to the NFV and SV groups. All NFV cases clinically exhibited AOS (“effortful speech”) and 5 patients (case 20-23 and 31) also displayed clinically relevant single-word comprehension deficits early in the disease and would also fit the more recently described criteria for the “mixed variant” (Mesulam et al., 2014; Schaeverbeke et al., 2018). It is of note that this subtype was described after enrollment for our study started. Hence, it is possible that some NFV enrolled before 2014 may also meet the criteria for the “mixed variant” (5 NFV were recruited before 2014). All patients received a volumetric MRI scan and 7 NFV and 7 SV DTI imaging. Twenty-nine healthy controls (15 male, age range 51-76, education range 9-22 years) performed the psychoacoustic tasks, 24 received volumetric MRI and 20 DTI imaging. Hearing sensitivity was measured in all participants using a clinical Bekesy-type audiometer for frequencies of 0.25, 0.5, 1, 2, 4 and 8kHz, on the left and right ear. Impaired pure-tone perception has been observed in NFV (Hardy et al., 2019), but here we included only participants able to detect stimuli of up to 1000 Hz below a hearing level of 30 dB on at least one side (Fig 6). Controls, NFV and SV were not significantly different in terms of age, gender, education or better-ear mean score (one-way ANOVA all P>0.136).

**Figure 6:**
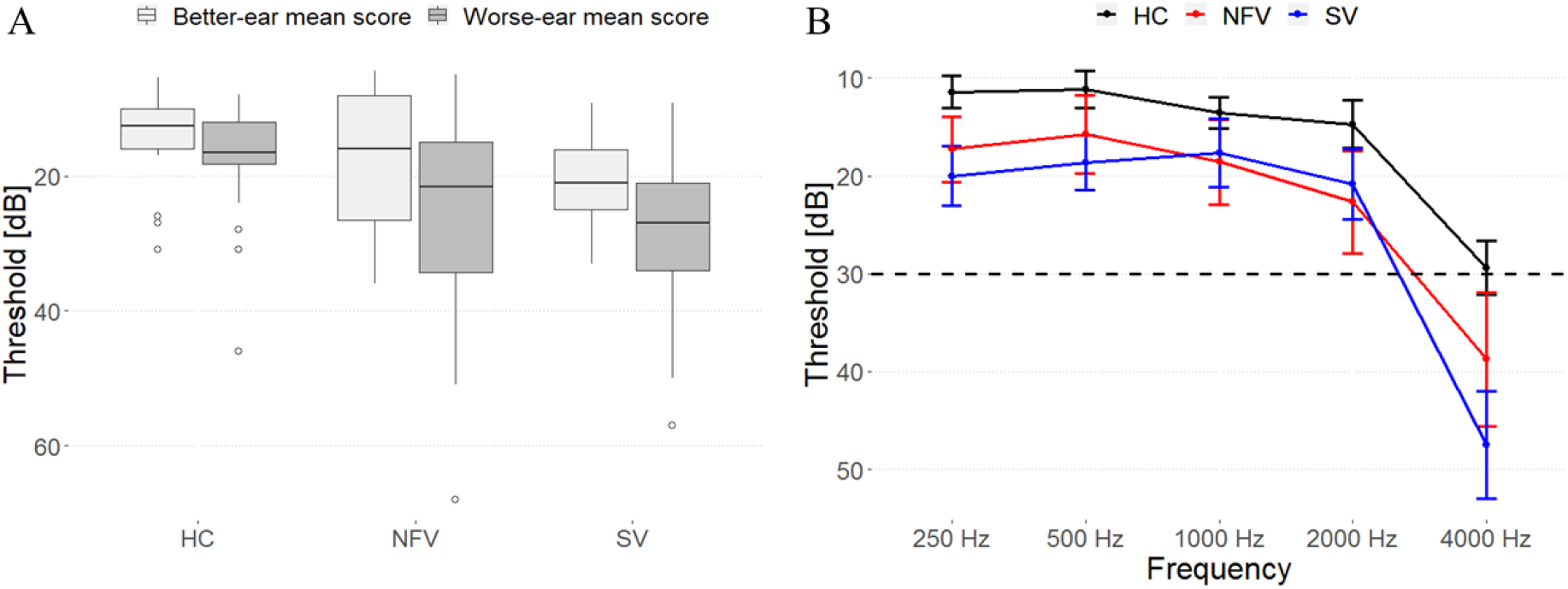
Pure-tone audiograms of all participants. A) Mean composite ear and frequency score (250-4000 Hz) data for each participant group. B) Mean thresholds (and standard error of the mean) for detection of tones at frequencies of 250, 500, 1000, 2000 and 4000 Hz for each participant group.

### Behavioral testing

Confrontation naming was tested using the Boston Naming test (Dutch norms). Non-verbal executive functioning was evaluated using Raven’s Coloured Progressive Matrices. Speech repetition was assessed using the Akense Afasie test. To assess AOS, the Diagnostisch Instrument voor Apraxie van de Spraak (DIAS) was added when it became available (for this reason it was not performed in 4/12 cases). The DIAS consists of vowel and consonant repetition (15 trials each) and diadochokinesis testing. During the latter task, the examiner first reads three successive alternating syllables aloud, e.g., “pa ta ka” and asks the patient to repeat these. If successful, he/she was asked to repeat it as many times as possible during a period of 8 s. The diadochokinesis severity score is the sum of correctly repeated syllables across trials. Grammaticality was assessed using the auditory sentence comprehension test of the Werkwoorden en Zinnen Test (WEZT), consisting of 40 sentence-picture matching trials with active or passive sentences containing possible role reversal (e.g. “the horse was kicked by the cow”).

### Connected speech analysis

To obtain a measure of speech rhythm, we determined the normalized pairwise variability index (PVI) in connected speech samples using Praat 6.1.02. The samples consisted of a 2-minute “Cookie Theft Scene” description (20 controls, 11 NFV, 9 SV). For every participant, the median PVI (Ballard et al., 2014) was determined for polysyllabic words with a strong-weak stress pattern (e.g. COO-kie) and for words with a weak-strong stress pattern (e.g. out-DA-ted). PVI was calculated following the procedure outlined in Ballard et al. (2014) and Duffy et al. (2017), equaling 100 x (d1 - d2)/ [(d1 + d2)/2], where d1 and d2 are the durations of the first and second vowel. Normalization corrects for a difference in speech rates. PVI is a marker of the suprasegmental timing of speech. PVI values closer to zero are consistent with relatively equal stress between the first two vowels of a word (Ballard et al., 2014). Additionally, speech rate was calculated for each sample, equaling the number of utterances per minute.

### Psychoacoustic tasks

Testing consisted of four pre-existing tasks (r1-r4) (Grube et al., 2016) (Fig 1A, supplementary audio files). The tasks followed a two-alternative forced-choice algorithm. Participants responded verbally or by pointing to a graphical scheme. Instructions, verbally and graphically, were repeated until the participant understood the task. Five practice trials were repeated until five consecutive correct responses were recorded, and if needed, instructions were repeated and the nature of the errors was explained. If the participant indicated during the test phase that they had forgotten the instructions, then they were repeated, the practice trials run again and the test phase then restarted.

All tasks used 500Hz 100 ms pure tones and consisted of 50 trials. The tasks were based on a two-alternative forced-choice adaptive paradigm following a 2-down, 1-up algorithm (difficulty increasing after 2 consecutive correct responses and decreasing after every incorrect one). A larger step size was used up to the fourth reversal and after that a smaller one. The outcome measure was the threshold, calculated as the mean over the last six reversals measured with the small step size, estimating the 70.9%-correct point of the psychometric function (Levitt, 1971). The difference between the target and distractor was varied as a relative proportion of the duration or tempo of the reference. The ‘Single time-interval duration discrimination’ task (r1) required participants to indicate which of two tone pairs contained the ‘longer gap’. Initially, the target was longer by 90% of the reference inter-onset-interval (depending on the trial, between 300 and 600 ms), and adaptively adjusted in steps of 12% and 6%. In the ‘Isochrony deviation detection’ task (r2), participants were instructed to indicate which of two otherwise isochronous five-tone sequences contained a lengthening or ‘extra gap’. The reference sequence had an isochronous inter-onset-interval ranging from 300 to 600 ms. The target had one lengthened inter-onset-interval between the third and fourth tone. The initial default value of the lengthening was 60% of the inter-onset-interval, adaptively adjusted in steps of 6% and 2%. In both tasks (r1,r2), a local deviation is introduced to generate the target. As such, these tasks test the detection of lower-order differences in timing between consecutive tones. In the ‘Metrical pattern discrimination’ tasks (r3, r4), participants had to decide which of three rhythmic sequences (the second or the third) of seven tones sounded “different”, based on a distortion within the rhythm. The reference sequence had a strongly (r3) or a weakly (r4) metrical beat of four evoked by the temporal spacing of the tones over 16 time units. In the strongly metrical sequence, accented tones occurred every four units, in the weakly metrical sequence, two of those were silent (Grube and Griffiths, 2009). The default initial distortion in pattern (a change in the long compared to the short intervals) was 65%, adaptively adjusted in steps of 12% and 6%. Metrical pattern discrimination (r3,r4) requires processing of the higher-order temporal structure of the stimuli, since global deviations distributed across the sequence need to be detected. Typical syllable rates in Dutch (the native language of the participants) are 4-5 syllables/s (period 200 – 250ms) which is close to the tempi used in our tasks.

### Statistical analysis

The analysis of the speech rhythm and rate and the psychoacoustic tasks was identical to Grube et al. (2016). Depending on the distribution, outcome measures were log-transformed to allow for parametric analysis at the group level. At the individual level, each patient’s performance was analyzed in comparison to the group by using a modified Crawford *t*-test (Crawford and Garthwaite, 2007). For the comparison between each patient and the controls, to facilitate comparison between tasks and to enable Bonferroni correction, the exact P values (estimated percentiles) calculated according to Crawford and Garthwaite were transformed into normalized Z-scores using the standard normal cumulative distribution function. The significance threshold was set to a one-tailed significance level of P<0.05, Bonferroni-corrected for the number of tests (n = 3 for the speech measures, n = 4 for the psychoacoustic tasks, one-tailed since the a priori hypothesis is that NFV would perform worse). For NFV, SV and controls, we correlated PVI and the speech rate to the psychoacoustic tasks to test the link between rhythmic abilities and speech rhythm (significance level of P<0.05). We compared the psychoacoustic thresholds between NFV and SV using a Student’s t-test (one-tailed significance level of P<0.05, effect size: Cohen’s d with Hedges correction for small samples, R package effsize).

### Acquisition of MRI data

Twenty-three patients (12 NFV, 11 SV), and 24 controls received a high resolution T1-weighted structural MRI. All controls and 13 patients were scanned on a 3T Philips Intera system equipped with an 8-channel receive-only head coil (SENSitivity Encoding head coil). Ten patients were scanned on a 3T Philips Achieva dstream scanner equipped with a 32-channel head volume coil. An identical 3D turbo field echo sequence was used on both systems (coronal inversion recovery prepared 3D gradient-echo images, inversion time (TI) 900 ms, shot interval = 3000 ms, echo time (TE) = 4.6 ms, flip angle 8°, 182 slices, voxel size 0.98×0.98×1.2 mm^3^). The diffusion weighted images consisted of 45 directions of diffusion weighting with a b = 800 as well as 1 non-diffusion weighted image (B0), acquired in the axial plane, with isotropic voxel size of 2.2 mm, TR 9900 ms, TE 90 ms, flip angle 90°, fold over direction AP, fat shift direction A (anterior), in-plane parallel image acceleration (SENSE) factor 2.5.

### Deformation-based morphometry

DBM was performed using the CAT12 toolbox (http://www.neuro.uni-jena.de/cat), an extension of SPM12 (http://www.fil.ion.ucl.ac.uk/spm). Segmentation was performed in CAT12 using a default tissue probability map. Local adaptive segmentation was used at default strength (medium) and Diffeomorphic Anatomical Registration Through Exponentiated Lie Algebra (DARTEL) was used for registration to the default template (IXI555_MNI152). Voxel size for normalized images was set at 1.5 mm (isotropic) after internal resampling at 1mm. Local deformations were estimated using the Jacobian determinant, while ignoring the affine part of the deformation field. Thus, additional correction for total intracranial volume is not required (Gaser and Kurth, 2019). Images were smoothed using a 8 x 8 x 8 mm^3^ Gaussian kernel. Deformation fields of controls and both PPA groups were compared using a one-way between-subject ANOVA. Multiple linear regression was used to correlate tests (r1-r4) at the individual level to the deformation fields within each PPA subtype. Scanner type and age were introduced as nuisance variables in all analyses. Threshold of significance was set at voxel-level uncorrected P<0.001 and cluster-level FWE-corrected P<0.05 (Grube et al., 2016).

### Diffusion Tensor Imaging

Diffusion images were preprocessed and analyzed with MRTRIX3. The preprocessing pipeline included the following steps: first, the data were converted to MIF using mrconvert. Using dwidenoise, diffusion data were denoised; subject motion, and eddy current artefacts were also corrected for using dwidenoise (which relies on FSL eddy); following these two steps, the preprocessed diffusion data were bias-corrected with dwibiascorrect. The diffusion data were rigidly aligned to the subject’s T1-weighted volume space using Advanced Normalization Tools (ANTs) and tensor reorientation was performed. Fractional anisotropy (FA) and mean diffusivity (MD) were calculated in subject-space and normalized to MNI space. The calculated tensors were then used to perform a whole brain tractography using the probabilistic Tensor (Tensor_Prob), combined with anatomically constrained tractography with seeding along the grey/white matter interface, and 2 million streamlines to be selected (Jones, 2008). The whole brain tractogram was then segmented using volumes of interest (VOIs) acquired from the Freesurfer aparc+aseg parcellation (Desikan et al., 2006; Fischl et al., 2004). These VOIs were pars opercularis of the IFG and the superior frontal gyrus, specifically selecting the Aslant tract on diffusion MR data (Catani et al., 2013).

Freesurfer aparc+aseg parcellation was performed to obtain these subject-specific VOIs. For this reason preprocessing of T1-weighted structural MRIs was repeated using FMRIPREP (Esteban et al., 2019), a Nipype (Gorgolewski et al., 2011) based tool. T1-weighted volume was corrected for intensity non-uniformity using N4BiasFieldCorrection v2.1.0 (Tustison et al., 2010) and skull-stripped using antsBrainExtraction.sh v2.1.0 (using the OASIS template). Brain surfaces were reconstructed using recon-all (FreeSurfer v6.0.1 (Dale et al., 1999)), and the brain mask estimated previously was refined with a custom variation of the method to reconcile ANTs-derived and FreeSurfer-derived segmentations of the cortical gray matter (Klein et al., 2017). Spatial normalization to the ICBM 152 Nonlinear Asymmetrical template version 2009c was performed through nonlinear registration with the antsregistration tool of ANTs v2.1.0 using brain-extracted versions of both the T1-weighted structural MRI and the template. Brain tissue segmentation of cerebrospinal fluid, white matter and gray matter was performed on the brain-extracted T1-weighted structural MRI using fast (FSL v5.0.9).

Smoothed FA and MD maps were compared between controls, NFV and SV using a between-subject ANOVA (same as previous threshold). Scanner type, TIV and age were introduced as nuisance variables. A template for the left Aslant tract was generated for healthy controls using the 75% overlap threshold (Catani et al., 2013). FA and MD of the left Aslant tract were extracted for each patient by averaging values from all voxels included in this template (Catani et al., 2013). We compared the FA and MD between NFV and SV by means of a Student’s t-test (one-tailed P<0.05). FA and MD were correlated to rhythm discrimination performance within the NFV group to confirm the DBM findings (one-tailed P<0.05).

## Data Availability

The data that support the findings of this study are available from the corresponding author upon reasonable request.

## Data availability

The data that support the findings of this study are available upon reasonable request.

## Acknowledgements

The authors thank B. Bergmans, MD, PhD, Ch. Swinnen, MD, A. Sieben, MD, and Y.A. Pijnenburg, MD, PhD, for the referral of patients. We thank E. Luckett, MSc, for copyediting.

